# High Frequency and Unique Subtypes of Meningioma in Patients with BAP1 Tumor Predisposition Syndrome

**DOI:** 10.64898/2025.12.04.25341363

**Authors:** Kaylee A. Ramsey, Lindsey Byrne, Olivia B. Taylor, Amr Soliman, Emma Schreiner, Isabella Gray, Alicia Latham, Rania Sheikh, Saman S. Ahmadian, Russell R. Lonser, Joshua D. Palmer, Maria I. Carlo, Colleen M. Cebulla, Mohamed H. Abdel-Rahman

**Author notes:** Corresponding Author: 400W 12^th^ Ave, Columbus OH, 43210.

## Abstract

**Background:** *BAP1*-tumor predisposition syndrome (*BAP1*-TPDS) is associated with four main cancers: uveal melanoma, cutaneous melanoma, malignant mesothelioma, and renal cell carcinoma. However, there are additional cancers found more rarely in *BAP1*-TPDS patients. The aim of this study was to investigate the association, clinical, and pathologic characteristics of meningioma in *BAP1*-TPDS.

**Methods:** We conducted a retrospective chart review of meningiomas in two independent cohorts of patients with germline *BAP1* pathogenic or likely pathogenic (P/LP) variants at The Ohio State University Wexner Medical Center and at the Memorial Sloan Kettering Cancer Center from October 1^st^, 2010 date to April 21^st^, 2025. Additionally, we conducted a literature review of meningioma case studies for individuals with germline *BAP1* (P/LP) variants.

**Results:** In a cohort of 237 subjects with *BAP1*-TPDS, we identified 6.8% (16/237) with history of meningiomas. The average age of diagnosis was 45.8 years (17-71). For patients with available pathology, 61.5% (8/13) of the tumors were grade II/III. Patients with available tumor tissue 83.3% (5/6) showed evidence of *BAP1* biallelic inactivation. Family history of meningioma was reported in 18.8% (3/16) of patients. Three cases of meningioma were identified during meningioma surveillance imaging. Published cases were consistent with the early age of onset, high-grade tumors, and clinical phenotype of tumors.

**Conclusions:** This study provides additional evidence that high-grade brain and spinal meningiomas are part of the clinical spectrum of *BAP1*-TPDS. Craniospinal imaging surveillance in the *BAP1*-TPDS population should be considered starting around puberty, enabling early detection and management for individuals with *BAP1*-TPDS.

**Key Points:**

- High frequency of patients in our BAP1-TPDS cohort had meningiomas (6.8%)
- BAP1-TPDS meningiomas had rhabdoid and papillary histologic subtypes and high WHO grades
- These patients were diagnosed at a younger age compared to the average population
- There was a female predominance of meningioma in BAP1-TPDS patients
- BAP1-TPDS related meningiomas are at risk for higher rates of local recurrence, particularly rhabdoid histology

**Importance of the Study:** BAP1-tumor predisposition syndrome (*BAP1*-TPDS) is a rare syndrome which is associated with four cancers: uveal melanoma, cutaneous melanoma, malignant mesothelioma, and renal cell carcinoma. However, our group has observed additional cancers in these patients, including meningioma. Here, we present a retrospective chart review of 16 patients with germline pathogenic *BAP1* variants and meningioma, representing 6.8% of our entire cohort of *BAP1*-TPDS cases. We report that BAP1-TPDS patients are at higher risk of developing high-grade meningiomas and are diagnosed at an earlier age than the general population. Three meningiomas and one meningothelial cyst were observed during surveillance imaging in asymptomatic patients. Patient and family cancer history included other *BAP1*-TPDS associated cancers, but only three patients had relatives with meningioma. Our findings indicate a need for routine craniospinal imaging in *BAP1*-TPDS patients, and surveillance should include patients without family history of meningioma and patients under the age of 30.

## Introduction

*BRCA1*-associated protein-1 (*BAP1*) is a tumor suppressor gene located on chromosome 3p21.1 and serves multiple roles in tumor biology. *BAP1* functions as a deubiquitinating enzyme, playing a crucial role in various cellular processes, including proliferation, DNA repair, differentiation, metabolism, and survival [1]. *BAP1* tumor predisposition syndrome (*BAP1*-TPDS) is an autosomal dominant hereditary cancer syndrome caused by heterozygous germline loss-of-function *BAP1* P/LP variants. [2–6]. The syndrome is associated with four main cancers including uveal melanoma, malignant mesothelioma, cutaneous melanoma, and renal cell carcinoma [3, 7, 8]. Affected individuals are also at risk of developing cutaneous BAP1-inactivated melanocytic tumors (BIMT), previously described as atypical Spitz tumors. The complete clinical phenotype of *BAP1*-TPDS has not yet been fully characterized and there is ongoing research suggesting the association of several other tumors [4, 6, 8–10]. Proper characterization of the clinical phenotype of the *BAP1*-TPDS is crucial for establishing management guidelines.

In the general adult population, total meningioma incidence is estimated to be approximately 10.82 per 100,000 (0.011%) [11]. Meningioma is the most common primary brain tumor in adults and displays a wide range of clinical and pathological features [12]. Although over 70% of meningiomas are regarded as benign, 5% -20% of patients with meningioma experience recurrences despite receiving maximal standard of care treatments [13, 14]. Higher grade meningiomas (II and III) account for 13-16% of meningiomas and are associated with higher rates of recurrence, up to 80% even after complete surgical resection, presenting significant treatment challenges [11, 15]. These tumors frequently lead to more aggressive symptoms and morbidity requiring repeated surgeries and radiotherapy which can substantially impact both quality of life and life expectancy.

Recent studies have identified *BAP1* mutations as a rare but important molecular alteration in meningiomas, potentially associated with poor clinical outcomes such as aggressive symptoms, recurrences, and shortened survival in some patients [16]. Though uncommon, loss of BAP1 function—due to either somatic or germline mutations—has been observed in a subset of meningiomas, particularly those with rhabdoid and/or papillary histologic features [16, 18–21]. Rhabdoid and papillary meningiomas are rare subtypes, each accounting for less than 1% of all meningiomas. The diagnosis of these rare subtypes is complicated by their heterogeneity and low incidence, which can lead to variability in histopathologic interpretation and subsequently impact decisions regarding adjuvant therapy [22]. Importantly, preclinical studies suggest that *BAP1*-deficient tumors may be sensitive to targeted therapies, including Enhancer of Zeste Homolog 2 (EZH2) inhibitors and Poly (ADP-ribose) polymerase 1 (PARP1) inhibitors [23, 24] and currently several clinical trials are ongoing [25]. As such, elucidating the genetic drivers—particularly *BAP1* alterations in high-grade meningiomas could meaningfully improve diagnostic precision, prognostic assessment, and treatment strategies for affected patients.

Both The Ohio State University Wexner Medical Center (OSUWMC) and Memorial Sloan Kettering Cancer Center (MSKCC) have specialized BAP1-TPDS clinical programs which incorporate standardized screening protocols for related BAP1 cancers. Currently, the National Comprehensive Cancer Network (NCCN) recommends screening for renal cell carcinoma with MRI abdomen in patients with *BAP1*-TPDS every 2 years beginning at age 30y [26]. Similarly, our OSU clinic recommends brain and spinal cord craniospinal MRI every 2 years beginning at age 30y for meningioma surveillance.

The goals of this study were to assess the evidence for association of meningiomas and the clinical phenotype of these tumors in the *BAP1*-TPDS. Our results provide strong evidence that high-grade meningiomas are part of the clinical spectrum of *BAP1*-TPDS.

## Methods

### Retrospective Chart Review

Our study integrated retrospective chart reviews from two institutional cohorts, OSUWMC and the MSKCC under IRB-approved protocols from October 1^st^, 2010 to April 21^st^, 2025. At OSUWMC, we included all patients with germline *BAP1* P/LP variants diagnosed, their family members, as well as subjects enrolled in the OSU *BAP1*-TPDS registry, “Frequency and Clinical Phenotype of BAP1 Hereditary Predisposition Syndrome” (NCT04792463). Individuals determined to be obligate carriers were included. Pedigrees were reviewed for reported cases of meningioma, and medical records were obtained to review the pathology. For tumors with available archival material (total six tumors) the pathology was reviewed by an expert neuropathologist to ensure classification, and the expression of BAP1 and proliferative marker Ki67 were assessed. Immunohistochemistry (IHC) was carried out at the OSUWMC Pathology Department, using a prediluted *BAP1* mouse monoclonal antibody (BioSB, clone BSB-109) and Ki-67 antibody. Pretreatment was carried out using Dako Flex High solution for 30 min and detection using the Flex Detection Kit for 20 min using the Omnis autostainer (Agilent, Dako).

At MSKCC, a retrospective chart review was conducted with all patients with germline BAP1 P/LP variants who had genetic consultation at the institution’s Clinical Genetics Service or underwent genetic testing through MSK-IMPACT. All patients were assigned a study ID code and clinical data regarding the patient’s age, sex, clinical presentation, and meningioma characteristics including pathology were obtained from the records.

### Literature Review

A review of published literature was conducted, focusing on identifying documented cases of germline *BAP1* P/LP variants in meningiomas (Figure 3). Published articles in PubMed and Scopus were reviewed for cases of patients with meningiomas and germline BAP1 P/LP variants to establish an association between BAP1-TPDS and meningiomas. Meningioma morphology, WHO grade, BAP1 P/LP variants, and clinical characteristics were recorded. It is important to note that two of the patients discussed in the literature review also belong to the OSUWMC institutional cohort.

## Results

### Demographic characteristics

The OSUWMC cohort had 13/195 cases while the MSKCC had 3/42 diagnosed with meningioma with a total of 16/237 in patients with germline *BAP1* P/LP variants (Table 1). These cases included 13 with confirmed pathological diagnosis and 3 cases with radiologically presumed or other diagnoses. One patient had a meningothelial cyst found along the spinal meninges a few years following the diagnosis and resection of her initial cranial meningioma. This patient and her daughter have been described previously [27, 28]. The mean age at presentation in this cohort is 45.1 years old (median = 46). Notably, the mean age at presentation for meningioma in the general population is 66 years old [11]. 75% of patients were females while only 25% patients were males. While most tumors were identified in symptomatic patients, three asymptomatic patients had tumors found during surveillance imaging for their known BAP1 P/LP variant status. During pedigree review, we identified an obligate carrier with a reported unspecified brain tumor, medical chart history for this deceased individual could not be collected to confirm diagnosis, therefore this patient was not included in the meningioma totals.

In the OSUWMC cohort, two patients with meningioma were related (mother and daughter) (Table 1), however, having a family member with meningioma was uncommon. Only one other patient, besides the mother and daughter (3/16), reported having a 1^st^ or 2^nd^ degree relative with meningioma. However, family history of other *BAP1* related cancers was typical with 81.3% (13/16) patients reported having one or more 1^st^ or 2^nd^ degree relatives with cancers associated with BAP1-TPDS, including cutaneous melanoma (7/16), uveal melanoma (5/16), mesothelioma (5/16), kidney cancer (4/16), basal cell carcinoma (3/16), and other cancers (Table 1). Among the patients with meningioma, 62.5% (10/16) had a separate primary tumor including kidney cancer (5/16), basal cell carcinomas (3/16), mesothelioma (2/16), breast cancer (1/16), cholangiocarcinoma (1/16), non-small cell lung cancer (1/16), uveal melanoma (1/16), and neuroendocrine tumor (1/16).

### Tumor Characteristics and Histopathology

A majority of meningiomas, 81.3%, were found in the cranium (13/16); 1 was found along the spinal meninges, and 2 were of unknown primary sites (Table 1). Twenty-three percent (3/16) meningiomas were radiographically presumed or otherwise did not have available pathology. For the pathology confirmed meningiomas, 38.5% (5/13) had rhabdoid morphology, 30.8% (4/13) had atypical pathology with unspecified subtype, 7.7% (1/13) had papillary, 7.7% (1/13) had both papillary and rhabdoid, and 7.7% (1/13) had transitional/mixed morphology (Figure 1, Figure 2). The morphology was not available for one patient (Table 1). Furthermore, we identified cystic lesions of the meninges in one patient, OSU-3, who was diagnosed with a brain meningioma and later had a cystic recurrence of her after surgical resection and adjuvant radiotherapy. During routine screening of her brain and spine, she was later found to have a meningothelial cyst at C7-C8 separate from the initial brain meningioma. WHO grading was conducted in 10 tumors, 20% (2/10) were WHO grade I, 40% (4/10) were grade II, 40% (4/10) were grade III. A total of 80% (8/10) of the tumors were considered high-grade. We were unable to confirm WHO grade for 6 of the tumors. Local recurrence of previously treated tumors was reported in 3 patients, two of which had rhabdoid pathology. The third recurrence was noted to be a cystic recurrence.

**Figure 1.**
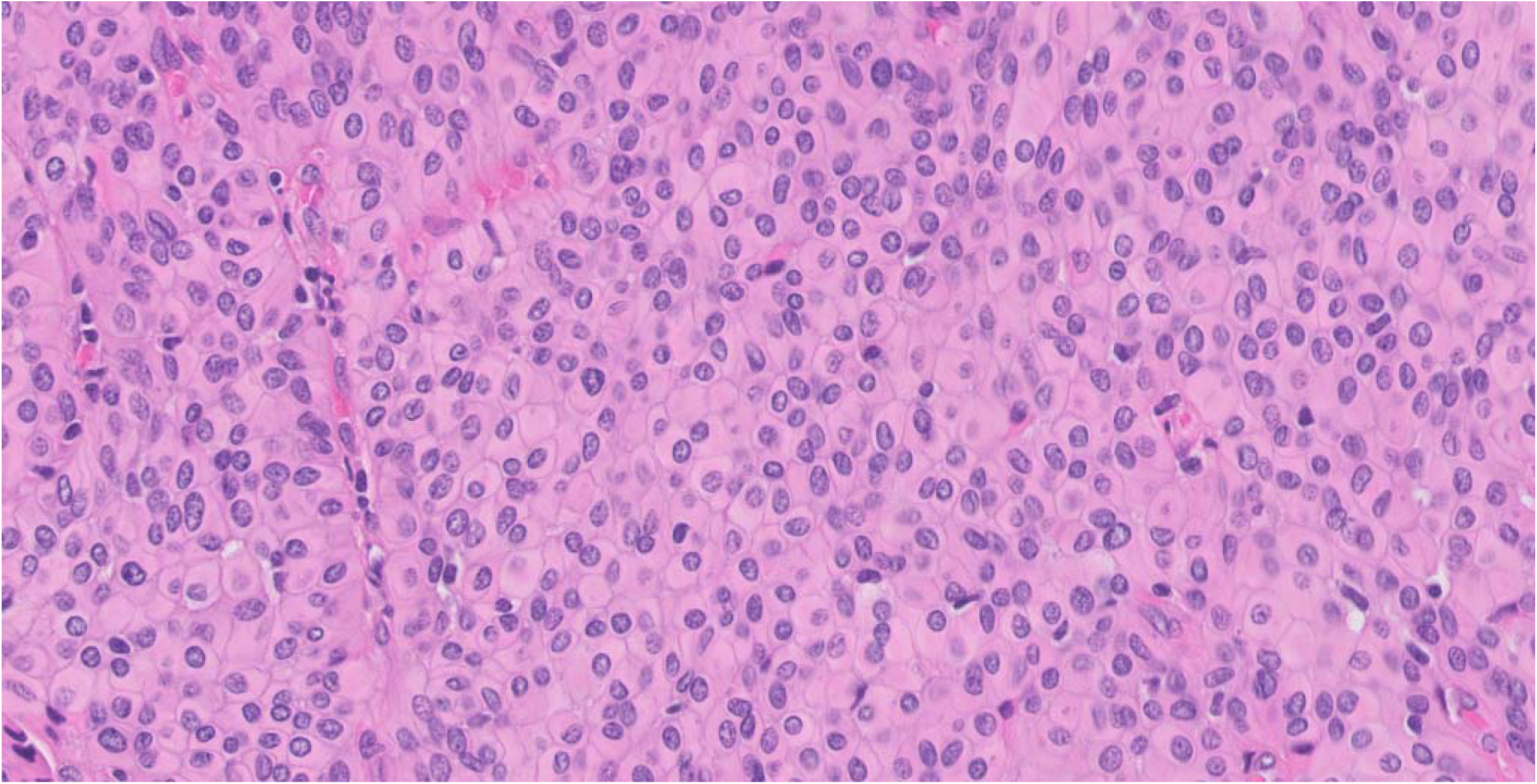
40x magnification Rhabdoid Meningioma (OSU-2)

**Figure 2.**
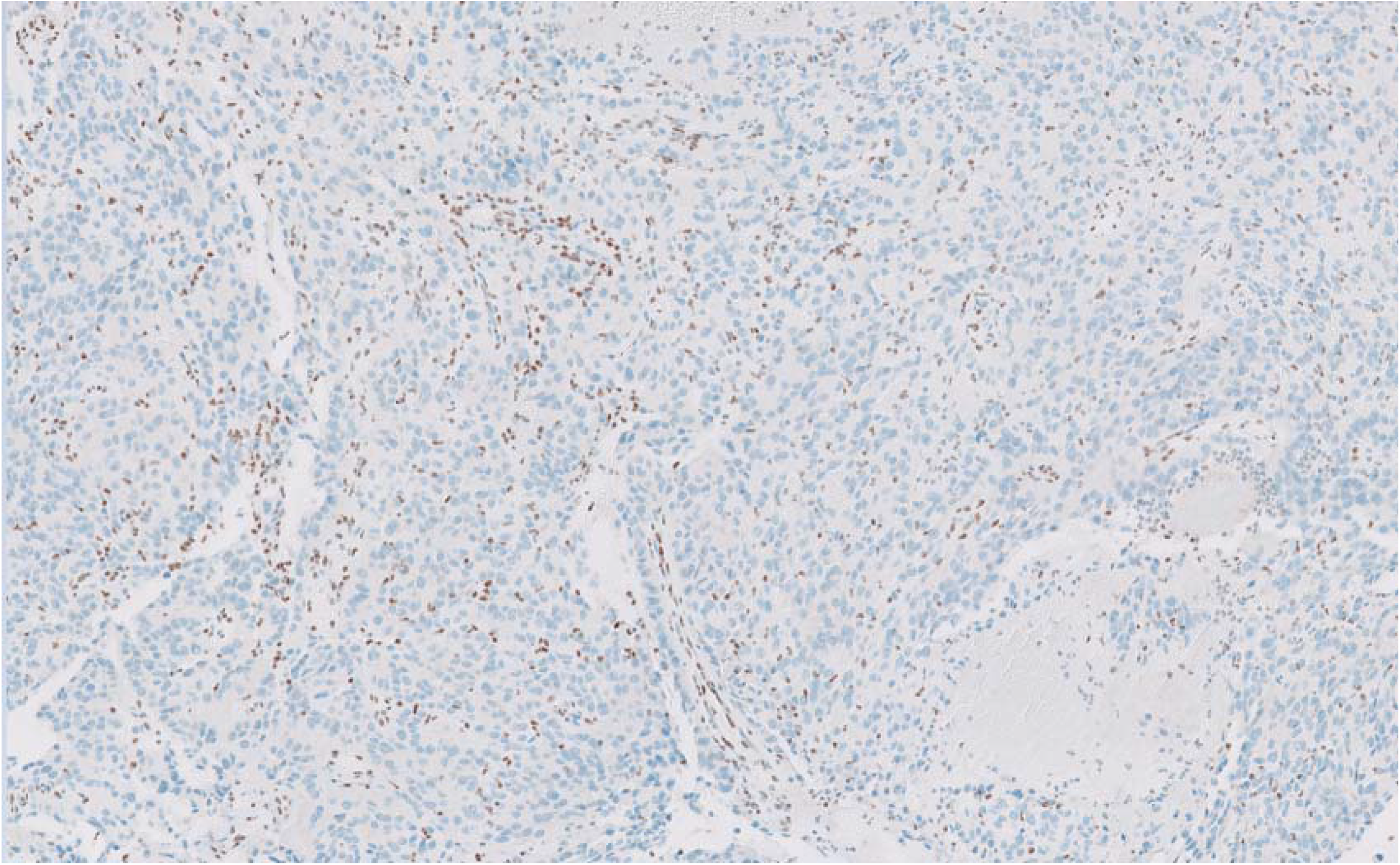
10x Magnification of WHO Grade I Rhabdoid/Papillary Meningioma with BAP1 Loss (OSU-3)

**Figure 3.**
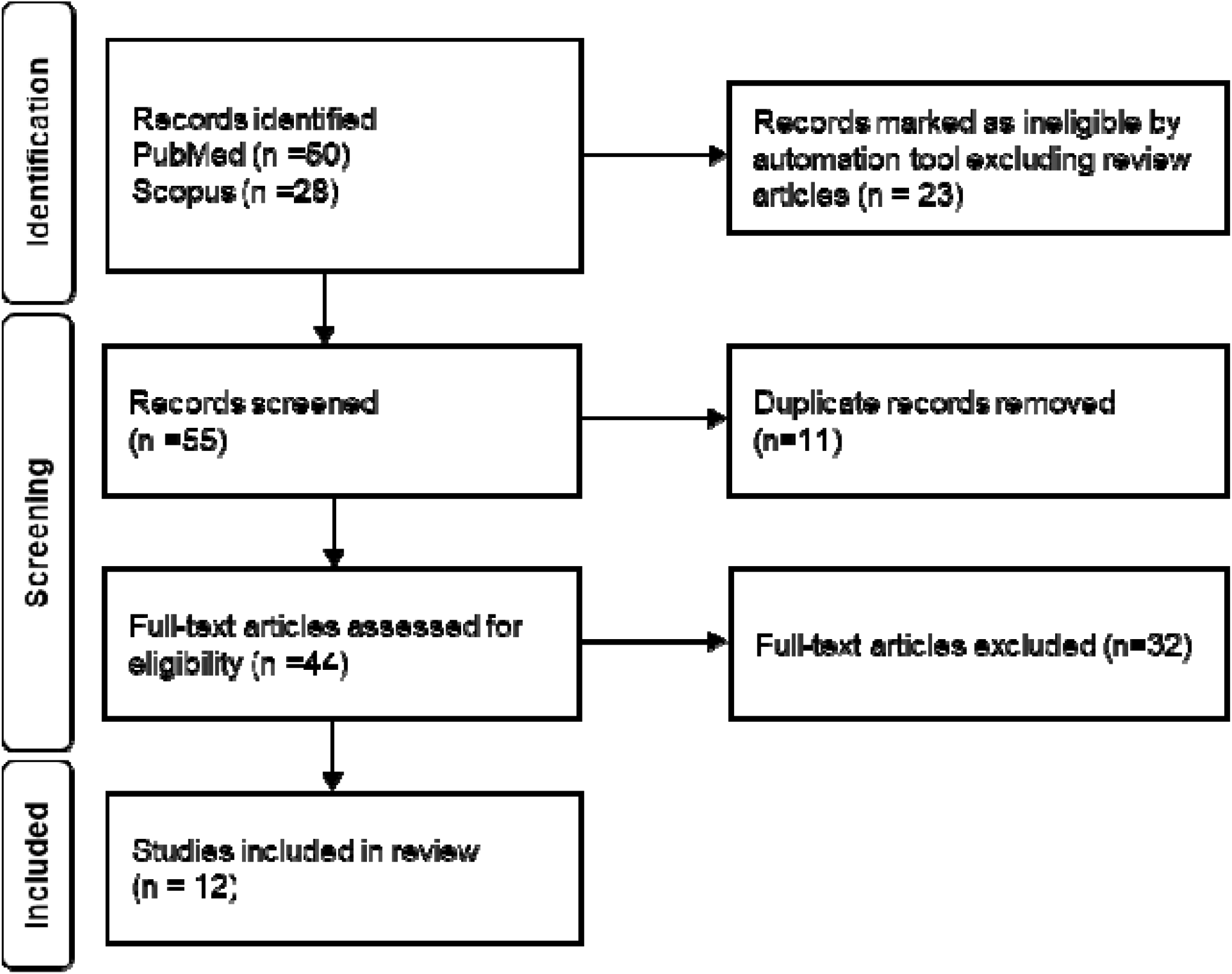
Review of Published Cases of Meningioma in patients with BAP1 Germline Pathogenic Variants

Tumor tissue was collected and re-analyzed for six patients (OSU-1, OSU-2, OSU-2, OSU-4, OSU-5, OSU-9) enrolled in the OSUWMC study to confirm the pathological characteristics. The proliferative index for these patients ranged from 2% to 15% (Table 2). For three patients, the mitotic count was less than 4 mitoses per 1.6 mm^2^. BAP1 loss was confirmed in 5 patients from the OSU cohort; BAP1 expression was preserved in the tumor tissue of one patient (OSU-9). Clinical pathology molecular tumor testing was only conducted in a subset of tumors. NF2 or TRAF7 alterations, which are common in some meningioma subtypes, were only tested in 3 tumors and were not detected. One patient (OSU-9) tested negative for CDKN2A.

### Literature Review

The comprehensive literature review aimed to ascertain the prevalence and characteristics of meningiomas lacking *BAP1* expression in published case reports. We identified a total of 13 patients with BAP1 P/LP variants and pathology confirmed meningioma (Table 3, Figure 1, Figure 2). Consistent with our cohort, the mean age at diagnosis was 46.7 years old (median= 56). 84.6% (11/13) of patients were females, 7.8% (1/13) of patients were male, and the gender of 7.8% (1/13) patients was not reported. Six patients were noted to have clinical symptoms that led to the discovery of their lesions, whereas the others were not specified.

76.9% (10/13) patients had intracranial meningiomas. The location of the meningioma was not specified in 23.1% (3/13) patients. Of the 10 patients with reported histopathology, 60% (6/10) were found to have meningiomas with predominantly rhabdoid morphology, 20% (2/10) patients were found to have a meningioma with papillary morphology, and 20% (2/10) patients were found to have both rhabdoid and papillary morphology [20, 28–32]. In publications where mitotic activity was measured in tumor sections, the mitotic range varied from very low overall (1 per 10 high-power fields [HPFs]) to very high (15 per 10 HPFs) [20, 29, 33]. Similarly, in publications where Ki67 antibody immunostaining was applied to tumor sections, the proliferative index ranged from 3%-15% [20, 33].

## Discussion

Recognizing the need for meningioma screening in individuals with germline *BAP1* P/LP mutations is an emerging priority in the management of *BAP1*-TPDS. Most meningiomas are slow-growing and benign [34]. However, recent case studies have reported that meningiomas with somatic and germline *BAP1* loss are particularly aggressive, and are associated with significant morbidity including seizure, focal neurologic deficits, altered mental status, weakness, and hydrocephalus [18–21, 29, 35]. Our study highlights the high prevalence of aggressive high-grade meningiomas in a subset of patients with germline *BAP1* P/LP variants. This observation prompts a re-evaluation of the tumor spectrum associated with *BAP1*-TPDS and underscores the potential importance of including meningiomas within the surveillance spectrum for these individuals.

### Frequency of meningiomas in BAP1-TPDS

Based on early data, our group’s prior estimated frequency of meningioma in *BAP1*-TPDS patients was 1.7% [17]. In the current study, evaluating both the OSUWMC and MSKCC cohorts, the frequency of meningioma in BAP1-TPDS was 6.7% (13/195) and 7.1% (3/42), respectively, with a combined frequency of 6.8% (16/237) supporting the high frequency in our previous study [8].

Collectively, these data represent a much higher prevalence among BAP1-TPDS patients than previously thought and further supports the published literature of increased incidence rates.

### Unique High Grade Histopathology Subtypes of BAP1-TPDS Associated Meningiomas

Meningiomas associated with *BAP1*-TPDS have most commonly been found to have rhabdoid morphology. Interestingly, in our cohort, there were patients found to have papillary, rhabdoid, or a combination of both rhabdoid and papillary morphologies. These data are further supported by studies by Shankar et al, in which 2 cases of germline BAP1-mutated meningiomas were found to have both papillary and rhabdoid morphology [29]. A recent report by Sievers et al found that 81% of somatic BAP1-altered meningiomas in their cohort had predominantly rhabdoid morphology but also reported papillary components, less commonly meningothelial and other mixed morphologies [36]. Rhabdoid and papillary morphology are both unique subtypes which usually are associated with higher grade features [37]. However, the 2021 WHO Classification of CNS Tumors no longer classifies meningiomas with rhabdoid and papillary subtype as high-grade tumors solely based on architecture and requires additional high-grade features for the designation of WHO grade 3 [12]. Even in the absence of these higher-grade features, BAP1 loss in these tumors is associated with more aggressive behavior including reduced time to recurrence, warranting close follow-up [38]. This suggests that mutations in BAP1 may not be limited to just rhabdoid morphology, and therefore the phenotype of meningioma in *BAP1*-TPDS should be expanded to include other subtypes including papillary, transitional and meningothelial [36]. Additionally, the loss of BAP1 in meningioma may be a predictor of aggressive clinical course irrespective of other high-grade. The incorporation of BAP1 in meningioma molecular testing is crucial for not only diagnosis and grading, but as predictor of aggressive behavior[36]. Given the rarity of BAP1-mutated meningiomas and the relative high frequency of germline alterations in these cases [16], reflex germline testing is recommended.

### Meningioma Treatment and Recurrence

In cases where the tumor is accessible, gross total resection followed by radiotherapy is the preferred primary treatment course for grade II or III meningiomas [39]. In the current study, all patients with available surgical and pathological data had undergone primary surgical resection and some received adjuvant radiation. Three patients had a local recurrence (OSU-2, OSU-3, and OSU-7; Table 1) demonstrating an incidence of 23.1% (3/13). One of our patients who was diagnosed with a intracranial meningioma was found to have a cystic recurrence of meningioma after surgical resection and adjuvant radiotherapy (patient OSU-3). She later was found to have a meningothelial cyst at C7-C8 during routine screening, which had not been described in earlier case reports of her and her daughter [27, 28]. In patient OSU-7, the initial tumor grade is unknown, however she underwent resection of her local recurrence, and the final WHO grade of this tumor was Grade III. In the literature review, patients with higher grade meningiomas had multiple recurrences [20, 29, 32, 33]. One patient was reported as having a metastasis which was distant from the initial meningioma [33]. The spectrum of disease associated with BAP1-TPDS may include risk of recurrence or of cystic lesions of the meninges. Further work is needed to study recurrence rates and best treatment options for BAP1-inactivated meningiomas.

### Screening recommendations for BAP1-TPDS patients

Standardizing screening for the *BAP1*-TPDS patient population is important; absence of formalized screening guidelines could lead to higher risk of delayed diagnosis and associated morbidity in patients with *BAP1*-TPDS. Our study showed that *BAP1*-TPDS patients are at a higher risk of developing meningioma which are diagnosed at an earlier age than the general population, emphasizing a need for standardized meningioma screening. Only 23.1% (n=3/13) patients from the OSU cohort reported having a 1^st^ or 2^nd^ degree relative with meningioma, indicating that their meningioma did not involve familial clustering. Many patients did have personal cancer history, and most also had a family history of cancers associated with *BAP1*-TPDS. Our results supports regular screening for meningioma in *BAP1*-TPDS patients irrespective of the family history of meningiomas.

Other cancers associated with *BAP1*-TPDS have screening guidelines: for example, to detect uveal melanoma, patients are recommended to have a yearly dilated eye exam beginning at age 11, which is 5 years earlier than the youngest patient diagnosed with uveal melanoma [40]. Remarkably, 25% of the cases in our cohort (4/16) and in the literature review (3/12) were diagnosed with meningioma prior to age 30 and as young as age 17. These findings suggest that biennial craniospinal imaging should begin prior to age 30, perhaps around puberty. If suspicious areas are noted on MRI, GA-68 DOTATATE PET scans are an option to help evaluate areas concerning for meningioma, as this technology can find small and multifocal meningiomas [41]. Clinicians should also take into consideration the psychological impact of screening, healthcare costs, and potential implications of detecting incidental findings.

### Study limitations

The OSUWMC cohort is enriched in patients with uveal melanoma and patients with personal/family cancer histories consistent with *BAP1*-TPDS, whereas MSKCC accrues patients with diverse cancers. Further research may allow for more robust data sets and more diverse manifestations of *BAP1*-TPDS to be detected and further refine our understanding of disease prevalence. In addition, there was a lack of clinical follow-up information on some patients and only part of the cohort was screened for meningioma with standardized craniospinal imaging protocols. Expanded screening data will be captured in a future publication. Finally, some of the lesions in our participants are radiographically presumed. Without tissue diagnosis, we could not confirm the etiology of the lesions with complete certainty. This also prevents us from ascertaining details regarding histopathologic cell type and BAP1 loss in the lesions.

## Conclusion

In conclusion, our findings underscore the value of considering meningioma screening in individuals with germline *BAP1* mutations. We observed a 6.8% prevalence of meningioma in two large cohorts of *BAP1*-TPDS patients. By utilizing a proactive approach to surveillance imaging, we allow for the potential for early detection, timely intervention, and personalized treatment strategies with the chance to improve outcomes for these individuals with *BAP1*-TPDS who have predisposition to the development of meningioma. Continued research into the clinical and molecular characteristics of meningiomas in the context of *BAP1*-TPDS will further guide the development of effective surveillance strategies and personalized therapeutic interventions for these individuals.

## Ethics Declarations

### Funding

National Institutes of Health R01-CA255323 (PI: Abdel-Rahman); Patti Blow Research Fund in Ophthalmology; Ohio Lions Eye Research Fund; OSU Cancer Center Core Grant 2P30CA016058-40; National Center For Advancing Translational Sciences 8UL1TR000090-05; OSU Vision Sciences Research Core Program P30EY032857; RPB New Chair Challenge Grant.

MSKCC-awarded grants: NIH K08-CA272960-02 (PI: Carlo); Kidney Cancer Association Trailblazer Award; NIH P30-CA008748.

### Conflict of Interest

The authors of this manuscript have no conflicts of interest to disclose.

## Acknowledgements

We would like to thank Shirley Ong, MD, Daniel Prevedello, MD for patient care.

## Authorship

Conception and design: LB, CC, MAR, KR, MC

Data acquisition, analysis and interpretation: KR, ES, IG, OT, MC, SA, AL, RS, AS

Writing and review: KR, OT, MAR, SA

Editing: KR, OT, LB, SA, MAR, CC, MC, RL

Study Supervision: MAR, CC

## Data Availability

All original data generated in support of this research will be made available at reasonable request to the corresponding author.

## Notes

### Competing Interest Statement

The authors have declared no competing interest.

### Clinical Trial

NCT04792463

### Funding Statement

This study was funded by National Institutes of Health R01-CA255323 (PI: Abdel-Rahman); Patti Blow Research Fund in Ophthalmology; Ohio Lions Eye Research Fund; OSU Cancer Center Core Grant 2P30CA016058-40; National Center For Advancing Translational Sciences 8UL1TR000090-05; OSU Vision Sciences Research Core Program P30EY032857; RPB New Chair Challenge Grant.
Research conducted by authors at MSKCCC was funded by NIH K08-CA272960-02 (PI: Carlo); Kidney Cancer Association Trailblazer Award; NIH P30-CA008748.

### Author Declarations

IRB of Ohio State University gave ethical approval for this work.

