## Supplementary material for "High Frequency and Unique Subtypes of Meningioma in Patients with BAP1 Tumor Predisposition Syndrome": Tables 1, 2, & 3

**Table 1: Review of Published Cases of Meningioma in patients with BAP1 Germline Pathogenic Variants**

| **Patient ID** | **Age^†^** | **Sex** | **BAP1 Variant** | **Meningioma Subtype and WHO Grade** | **Location** | **Presentation** | **Intervention, follow up duration and Outcome** | **Recurrence** | **Personal Cancer History** | **Family Cancer History^§^** |
| --- | --- | --- | --- | --- | --- | --- | --- | --- | --- | --- |
| OSU-1 | 60-65 | F | c.799C>T, p.Gln267* | Mixed, grade N/A | Right parietal brain | N/A | Resection | Unknown | None | Uveal melanoma, benign meningioma, lung adenocarcinoma, mesothelioma (2), cutaneous melanoma, ovarian, kidney, esophageal, colon, stomach, testicular |
| OSU-2^‡^ | 20-25 | F | c.1777C>T, p.Gln593*  (daughter and mother) | Atypical, Rhabdoid, II | Extra-axial, prepontine cistern | Symptomatic | Resection and adjuvant radiotherapy for initial tumor, radiation to recurrence. Stable for 64 mos | Yes, at 73 mos | None | Atypical meningioma, lung, kidney |
| OSU-3^‡^ | 55-60 | F |  | 1) Atypical, Rhabdoid/  Papillary, II  2) Meningothelial cyst^┼┼^ | 1) Left jugular foramen  2) T8/9 | 1) Symptomatic  2) Asymptomatic, discovered on surveillance scans. | 1) Resection and adjuvant radiotherapy, persistent cystic component, monitored. 59 months, stable disease.  2) Resection, 31 months, stable disease. | Yes | Multiple Basal Cell Carcinomas |  |
| OSU-4 | 15-20 | M | c.1379C>G, p.Ser460* | Rhabdoid, II | C6/C7 | Symptomatic | N/A | Unknown | None | Skin (unknown type) (2), bladder |
| OSU-5 | 25-30 | F | c.877_878del, p.Pro293fs*13 | Rhabdoid, III | Fronto-parietal lobe | Symptomatic | Received adjuvant radiotherapy at time of resection, follow-up unknown | No | None | Brain, uveal melanoma, squamous cell carcinoma, uveal melanoma, breast |
| OSU-6 | 55-60 | F | c.660-2A>G | Atypical, subtype N/A, III | N/A | N/A | N/A | Unknown | None | Cutaneous melanoma, mesothelioma, breast, colon, uveal melanoma, multiple myeloma |
| OSU-7 | 40-45 | F | c.1717delC (p.Leu573pfs*3) | Rhabdoid, III | Right occipital lobe | Symptomatic | Initial treatment with radiotherapy, recurrence treated with surgical excision and adjuvant radiation | Yes, time frame unknown | None | Cholangiocarcinoma (2), multiple basal cell carcinomas, vulvar, cutaneous melanoma, breast, mesothelioma (3), colon (2) |
| OSU-8 | 45-50 | M | c.37+1G>T | Papillary, III | Anterior cranial fossa | Symptomatic | Resection, stable for 30 mos | No | Basal Cell Carcinoma | Cervical, skin (unknown type), breast |
| OSU-9 | 60-65 | M | EX12_3'UTRdel | N/A, II | Anterior clinoid and left cerebellar hemisphere | Asymptomatic, discovered on surveillance scans | Resection and radiotherapy to residual disease, 15 months, stable disease. | No | Renal Cell Carcinoma | Cutaneous melanoma, prostate, breast, pancreatic (3), throat, renal cell carcinoma (2) |
| OSU-10 | 60-65 | F | c.1891-1G>A | N/A | N/A | Symptomatic | N/A | Unknown | Renal Cell Carcinoma, pancreatic neuroendocrine cancer | Cutaneous melanoma, multiple basal cell carcinomas, breast (2), uterine |
| OSU-11 | 40-45 | F | c.37+1G>T | Atypical, I | N/A | Symptomatic | N/A | Unknown | Renal Cell Carcinoma | Breast (2), cutaneous melanoma, thyroid |
| OSU-12 | N/A | F | EX10_11del | Radiographically Presumed, grade N/A | Brain | Asymptomatic, discovered on surveillance scans | N/A | Unknown | Breast, Renal Cell Carcinoma, Cholangio- carcinoma | Uveal melanoma, breast, mesothelioma, cutaneous melanoma, renal cell carcinoma |
| OSU-13 | 15-20 | F | c.1153C>T, p.Arg385* | Radiographically Presumed, grade N/A | Brain | N/A | N/A | Unknown | Uveal melanoma | Lung (2), bladder, esophageal |
| MSKCC-14 | 30-35 | M | c.1675_1684delACAGGCCTGC | N/A | N/A | N/A | Resection | Unknown | Non-small cell lung, mesothelioma | Pancreatic, cutaneous melanoma |
| MSKCC-15 | 55-60 | F | c.1254T>A, p.Tyr418* | Atypical, grade N/A | Left frontoparietal region | Symptomatic | Resection | Unknown | Mesothelioma, kidney, breast | Bladder, peritoneal mesothelioma vs. ovarian carcinoma |
| MSKCC-16 | 70-75 | F | c.1203T>G,p.Tyr401* | Rhabdoid, I | Left orbital roof/floor of anterior cranial fossa | Asymptomatic, discovered on surveillance scans | Resection | Unknown | Basal cell carcinoma | Pancreatic, squamous cell carcinoma, basal cell carcinoma, uveal melanoma, brain, liver |

†Age in years at diagnosis. ‡Patient 2 is the daughter of Patient 3, they are included in Hu et al, 2022 [27] and Prasad et al, 2021 [28]. §1^st^ and 2^nd^ degree relatives were counted.

Acronyms: WHO, World Health Organization; OSU- Ohio State University; MSKCC, Memorial Sloan Kettering Cancer Center; N/A, not available. ^┼┼^ Meningothelial cyst was not included in the total meningioma numbers.

**Table 2. Pathology characteristics of Meningiomas from OSU Cohort**

| **Patient ID** | **Morphology Rhabdoid and Papillary** | **Brain Invasion** | **Ki67** | **Mitotic Count**  **(per 1.6 mm^2^)** | **NF2;TRAF7** | **BAP1 IHC/Genotyping** | **WHO Grade** |
| --- | --- | --- | --- | --- | --- | --- | --- |
| **OSU-1** | Both | N/A | N/A | N/A | N/A;N/A | Loss of nuclear expression/LOH | NA |
| **OSU-2** | Rhabdoid | Negative | 15% | < 4 (3 mitoses) | Neg;N/A | Loss of nuclear and cytoplasmic expression | II |
| **OSU-3** | Both | Negative | 10% | <4 (1 mitosis) | Neg;N/A | Loss of nuclear and cytoplasmic expression | II |
| **OSU-4** | Rhabdoid | Negative | 8% | 8 | N/A;N/A | Loss of nuclear and cytoplasmic expression | II |
| **OSU-5** | Rhabdoid | Negative | 8% | N/A | N/A;N/A | Loss of nuclear and cytoplasmic expression | III |
| **OSU-9** | Neither | Positive | 2% | <4 (1 mitosis) | Neg/Neg | Positive nuclear expression | II |

Abbreviations: OSU, Ohio State University; IHC, immunohistochemistry; WHO, World Health Organization; LOH, loss of heterozygosity; N/A, Not available

**Table 3: Literature Review of Published Cases of Meningioma in patients with BAP1 Germline Pathogenic Variants**

| **Reference** | **Age** | **Sex** | **BAP1 Variant(s)** | **Meningioma Subtype and WHO Grade** | **Location** | **Presentation** | **Evidence of biallelic inactivation** | **Intervention and outcome** | **Personal cancer history** | **Family cancer history** |
| --- | --- | --- | --- | --- | --- | --- | --- | --- | --- | --- |
| Shankar et al (2017)[26] | 55-60 | F | p.G220_ splice (exact mutation N/A) | Rhabdoid, III | Left frontal | N/A | Loss of nuclear BAP1 in tumor | Unknown treatment, lost to follow-up | Unknown | Mesothelioma |
|  | 50-55 | M | c.519T>G (p.Tyr173*) | Rhabdoid, III | Left parietal | N/A | Loss of nuclear BAP1 in tumor | Resection and adjuvant radiation, three recurrences | Unknown | Unknown |
| Ravanpay et al (2018)[20] | 10-15 | F | c.1174C>T, p.Gln392* | Rhabdoid, III | Right tentorium | Symptomatic | Loss of nuclear BAP1 in tumor | Resection and adjuvant radiation, multiple recurrences | None | Leukemia |
| Cheung et al (2015)[27] | NA | F | c.1938T>A, p.Tyr646* (sisters) | N/A | N/A | N/A | N/A | N/A | Mesothelioma (pleural), basal cell carcinoma | Mesothelioma (n=2), uveal melanoma (n=1), breast (n=1), cutaneous melanoma (n=2), basal cell carcinoma (n=1) |
|  | NA | F |  | N/A | N/A | N/A | N/A | N/A | Mesothelioma (peritoneal) |  |
| Hu et al (2022)[28] | 60-65 | F | c.778C>T (mother and daughter) | Radiographically presumed | Left frontoparietal convexity | Symptomatic | Loss of nuclear BAP1 in mesothelioma tumor | Deceased prior to pathologic confirmation | Multiple basal cell carcinomas, pleural mesothelioma | Mesothelioma (n=3), renal cell carcinoma (n=2), bladder (n=1), melanoma (n=1) |
|  | 40-45 | F |  | Papillary, III | Right sphenoid sinus. | Symptomatic | N/A | Resection, adjuvant radiation, deceased ~ 48 months after diagnosis | Pleural mesothelioma |  |
|  | 70-75 | F | c.1717delC | Rhabdoid, III | Adjacent to right sylvian fissure | Symptomatic | Loss of nuclear BAP1 in tumor | Resection, deceased < 15 months after resection | Pleural mesothelioma, multiple basal cell carcinomas, breast cancer, colon cancer, lung cancer | N/A |
|  | 20-25 | F | c.1777C>T, p.Gln593  (daughter and mother)^†^ | Rhabdoid, II | Extra-axial, prepontine cistern | Symptomatic | Loss of nuclear BAP1 (Table 2) | Resection and adjuvant radiotherapy,  stable disease | None | Mesothelioma (n=1), renal cell carcinoma (n=1) |
|  | 55-60 | F |  | N/A, Grade II^‡^ | Not specified (see Table 1) | Symptomatic | Loss of nuclear BAP1 (Table 2) | Not specified (see Table 1) | Not listed (see Table 1) |  |
| Manookian et al (2024)[29] | 15-20 | F | c.1478_1479delCA (p.Thr493Argfs*5) | Rhabdoid, II | Brain, unspecified | Incidental (MRI for post-concussive headache) | Copy number loss affecting entire chromosome 3 | N/A | N/A | N/A |
| Biczok et al (2023)[31] | 55-60 | M | c.118C>T (p.Gln40*) | Papillary, III | Intraventricular | N/A | Focal copy number loss | Resection, second tumor appeared after 12 months, intradural adjacent to L5 | Uveal melanoma, basal cell carcinoma | N/A |
| Pandithan et al (2022)[30] | 55-60 | F | Whole gene deletion | Rhabdoid, III | Sphenoid wing and temporal  convexity (separate tumors) | N/A | Loss of nuclear BAP1 in sister’s hepatoid tumor | Two separate recurrent tumors with multiple excisions | Uveal melanoma, basal cell carcinoma, mesothelioma | Basal cell carcinoma (n=5), hepatoid carcinoma of the pancreas (n=1), stomach (n=1), cutaneous melanoma (n=1) |

†Patient 4 and 5 from Hu et al are also included in our institutional cohort as OSU-2 and -3, and are described by Prasad et al, 2021 [28]. ‡At OSU, this tumor was classified as a WHO Grade I meningioma with papillary and rhabdoid features.

Acronyms: WHO, World Health Organization; N/A, not available
